# Pathways from Polygenic Risk to Suicidality: Effects of Alcohol Use Disorder and Childhood Adversity

**DOI:** 10.64898/2026.02.10.26345999

**Authors:** Victoria Wu, Xue-Jun Qin, Allison Ashley-Koch, Nathan A. Kimbrel, Joel Gelernter, Anna R. Docherty, Henry R. Kranzler, Richard Feinn, Christal N. Davis

## Abstract

**Background:** The prevalences of suicidal ideation (SI) and suicide attempt (SA) are influenced by genetic, behavioral, and environmental factors. Alcohol use disorder (AUD) and adverse childhood experiences (ACEs) may mediate or moderate genetic liability for suicidality.

**Methods:** Using data from 10,275 participants (43.8% female; 47.2% African-like genetic ancestry [AFR], 52.8% European-like genetic ancestry [EUR]), we tested whether polygenic scores (PGS) for SI and SA predicted lifetime SI or SA. We also evaluated whether alcohol use disorder (AUD) mediated these associations and whether adverse childhood experiences (ACEs) moderated the direct and indirect pathways.

**Results:** Although there were significant direct associations of the SA PGS with SA (AFR: b = 0.36, SE = 0.01; EUR: b = 0.17, SE = 0.01; both *p*s < 2e-16), the SI PGS did not predict SI (p > 0.55). AUD mediated SA genetic risk (average causal mediation effect (ACME): AFR = 0.01, 95% CI [0.01-0.01]; EUR = 0.02, 95% CI [0.01-0.02]; both *p*s < 2e-16). Moderation analyses indicated that indirect effects were attenuated by ACEs score (ΔACME: AFR = 0.02, p < 2e-16; EUR = 0.01, p = 0.03). There was neither mediation nor moderated mediation for SI.

**Conclusions:** Genetic liability to SA operates partly through AUD, particularly among individuals with lower childhood adversity. Under higher adversity, alternative pathways to SA likely predominate. These findings highlight the need to consider distinct etiological pathways to the development of suicidality and the relevance of AUD as a modifiable target for suicide prevention among individuals at high genetic liability.

## Introduction

Worldwide, suicide claims more than 700,000 lives annually and is the third leading cause of death among 15-29-year-olds.^1,2^ Despite considerable investment in prevention, U.S. suicide mortality rates have risen steadily over the past two decades.^3,4^ Addressing this trend requires greater clarity on the factors that contribute to risk across the spectrum of suicidal thoughts and behaviors. Definitions of suicidality span from passive death wishes to suicidal ideation (SI; i.e., thoughts of killing oneself), plans, and suicide attempts (SA).^5^ Although suicidality is often thought of as occurring in the context of psychiatric disorders, most individuals with a psychiatric disorder will never make a suicide attempt or die by suicide, underscoring the need to examine other risk factors.^6^ Thus, suicidality is not merely a symptom or consequence of psychiatric disorders like major depression, but a complex phenotype shaped by genetic, behavioral, and environmental factors.

Twin and family studies estimate the heritability of SI at 30%-50%, indicating the importance of genetic contributions.^7^ Genome-wide association studies (GWAS), in contrast, identify single-nucleotide polymorphism (SNP)-based heritability estimates of only 3-4%.^8^ For SA, heritability is somewhat higher at 40-55% in twin studies,^7^ with SNP-based heritability typically in the range of 4-7%.^9^ Although SI and SA are moderately genetically correlated (r_g_ ≈ 0.5-0.7), they share <50% of their genetic variance,^9^ which underscores the need to differentiate the traits etiologically. Polygenic scores (PGS), which aggregate small effects of common variants across the genome, enable the estimation of an individual’s genetic liability, with SI and SA PGS being associated with their respective traits.^10^ Thus, PGS are useful tools to elucidate genetic risk for suicidality, though their predictive utility is limited.

Among behavioral risk factors for suicidality, alcohol use disorder (AUD) is one of the most potent. Studies show ∼2-10-fold higher odds of SA for individuals with AUD compared to the general population.^11,12^ Alcohol use acutely elevates SA risk, with individuals having a 6-7-fold increase in SA odds within 24 hours of consumption.^13^ Supporting this acute risk, toxicology studies frequently detect alcohol in suicide decedents,^14^ and ∼40% of treatment-seeking AUD inpatients report a lifetime attempt.^15^ Population-based studies demonstrate that alcohol dependence also increases the odds of SI independent of mood disorders and other psychiatric comorbidities,^11^ while there is longitudinal evidence that problematic drinking predicts both the onset and persistence of SI.^16^ Moreover, acute intoxication may transiently increase suicidal thoughts, which often diminish once individuals are no longer intoxicated.^17^ The robust link between alcohol and suicidality reflects the impacts of reduced inhibitory control, heightened impulsivity, and narrowed attentional focus to proximal stressors (“alcohol myopia”), all of which may amplify suicidal thoughts and accelerate the transition from SI to SA.^18^

Adverse childhood experiences (ACEs), which include abuse, neglect, and household dysfunction, also show robust dose-dependent associations with SI, SA,^19,20^ and AUD.^21,22^ For example, in the landmark Centers for Disease Control (CDC)-Kaiser ACEs study of 17,337 U.S. adults, exposure to ≥4 ACEs conferred more than 10-fold odds of a lifetime SA compared to those with no ACEs exposure.^23^ Similar effects have been observed for SI.^19,20^ An individual’s cumulative ACEs burden predicts a wide range of mental health outcomes, including AUD.^24^ Unfortunately, ACEs are common, with approximately 60% of U.S. adults reporting at least one ACE.^25^

Genetic liability and environmental stressors such as ACEs may interact to influence risk for suicidality,^26,27^ in line with diathesis-stress or differential-susceptibility frameworks.^28,29^ The diathesis-stress model proposes that inherited vulnerabilities (e.g., higher PGS) may remain latent until activated by environmental stressors.^28^ Thus, in the context of ACEs, genetic effects on suicidality may be amplified. In contrast, the differential-susceptibility framework posits that some individuals (perhaps due to genetic variation) have greater environmental plasticity, such that their risk is magnified in adverse environments, with greater relative benefit in supportive environments.^29^ These models are both consistent with ACEs moderating the association between genetic risk and suicidality. AUD may also mediate the effect of genetic liability on outcomes (PGS → AUD → SI/SA), consistent with robust evidence linking AUD to suicide risk,^11–18^ and in line with prior work showing that alcohol dependence accounts for between 8.5% to 47.1% of the association between ACEs and SA.^30^

### Current Study and Hypotheses

To evaluate the associations among genetic risk, AUD, and suicidality outcomes, we used data from the Yale-Penn cohort, a large, deeply phenotyped sample enriched for individuals with substance use disorders. We have reported prior GWAS for alcohol dependence^31^ and suicide traits^32^ in this sample. First, in ancestry-stratified mediation models, we examined the association of PGS for SI and SA with their respective traits, with AUD as a mediator of the associations. Next, we examined ACEs as moderators of the direct and indirect paths. We hypothesized that (1) PGS would exert both direct and indirect effects on suicidality via AUD; (2) the mediating effects of AUD would be stronger for SA than SI, reflecting alcohol’s effects on impulsivity and disinhibition—features that are more tightly linked to SA than SI; and (3) ACEs would moderate direct and indirect effects of PGS, amplifying genetic risk in the context of high adversity, in line with diathesis-stress models.^19,20,23,28,29^

## Methods

### Participants and Procedures

Participants were from the Yale-Penn cohort (n = 10,275; 43.80% female; 47.21% African-like genetic ancestry [AFR], 52.79% European-like genetic ancestry [EUR]), a deeply phenotyped sample recruited for studies of the genetics of substance use disorders, which also included screened controls. Recruitment occurred at five U.S. sites through addiction treatment centers, psychiatric services, and community advertisements. All participants provided written informed consent, and the study received approval from the institutional review board at each site. Participants were interviewed with the Semi-Structured Assessment for Drug Dependence and Alcoholism (SSADDA), a computer-assisted interview that includes assessment of suicidality, AUD, demographics, and environmental factors.^33^ Participants also provided blood or saliva samples for DNA extraction and genome-wide genotyping.

### Phenotypes

Suicidality was assessed using two questions. Participants reported whether they had experienced lifetime SI (i.e., “Have you ever thought about killing yourself?”) and whether they had made a lifetime SA (i.e., “Have you ever tried to kill yourself?”). AUD was modeled as a latent factor based on responses to the 11 Diagnostic and Statistical Manual of Mental Disorders, version 5 (DSM-5) AUD criteria assessed via the SSADDA.^34^ Model fit for the AUD factor was good-to-excellent (CFI = 0.95; RMSEA = 0.07; SRMR = 0.03), with all 11 criteria loading significantly (standardized loadings from 0.52 to 0.78; see Supplementary Table S1). The resulting factor scores, standardized within ancestry group, served as mediators in all models.

ACEs were also modeled as a latent factor,^35^ using 10 dichotomized indicators reflecting adversity or a lack of protective factors before age 13: multiple main caregivers (≥3); multiple relocations (≥2); witnessing/experiencing violent crime, sexual abuse, or physical abuse; household substance use; household smoking; religious participation (reverse coded); caregiver relationship quality (reverse coded); and contact with relatives (reverse coded). All items loaded significantly (p < 0.001; see Supplementary Table S2), and residuals for household substance use and smoking were allowed to covary to improve fit. The resulting model fit was good (CFI = 0.92; RMSEA = 0.04; SRMR = 0.03).

### Genotyping, Imputation, and Polygenic Scores

Yale-Penn participants were genotyped on Illumina microarrays, and quality control procedures were applied before imputation to the Haplotype Reference Consortium (HRC) reference panel. Principal components (PCs) of genetic ancestry were derived using genome-wide data and the 1000 Genomes Project reference panels to adjust for population stratification.^36^ Including PCs in the models helps account for genetic differences related to historical migration patterns that could otherwise confound PGS associations. Additional details on these procedures can be found elsewhere.^37,38^

To estimate individuals’ genetic liability for suicidality, we calculated SI and SA polygenic scores (PGS) using PRS-CS software and summary statistics from large GWAS. PRS-CS uses a Bayesian approach to infer posterior effect sizes of SNPs.^39^ We used the default settings to estimate the global shrinkage parameter and set a random seed of 1 to ensure reproducibility. The SI PGS were calculated using summary statistics from an ancestry-matched GWAS of SI in the absence of SA or suicide death conducted in 99,814 cases and 512,567 controls from the Million Veteran Program.^8^ The SA PGS used summary statistics from an ancestry-matched GWAS meta-analysis of 43,871 cases and 915,025 controls.^40^ All PGS were standardized within genetic ancestry group.

### Statistical Analyses

Analyses were conducted in the *mediation* package in R (v4.3) within each genetic ancestry group. We tested whether the AUD factor mediated the association between the PGS for SI and SA and lifetime suicidality outcomes. The mediation analysis followed a standard two-model framework: one model estimating the effect of the PGS on the mediator, and a second estimating the effect of the mediator on the outcome while adjusting for the PGS. This approach separates the total PGS effect into its direct and indirect components. For each ancestry group, the following paths were estimated (Figure 1): (1) path a: a linear regression predicting the AUD factor from the suicidality PGS, which indicates the degree to which genetic liability for suicidality increases alcohol involvement; (2) path b: a logistic regression of suicidality outcome on the AUD factor, controlling for the PGS, which indicates how AUD affects suicidality risk independent of genetic liability; and (3) path c’: the direct association between the PGS and suicidality outcome, controlling for AUD.

**Figure 1.**
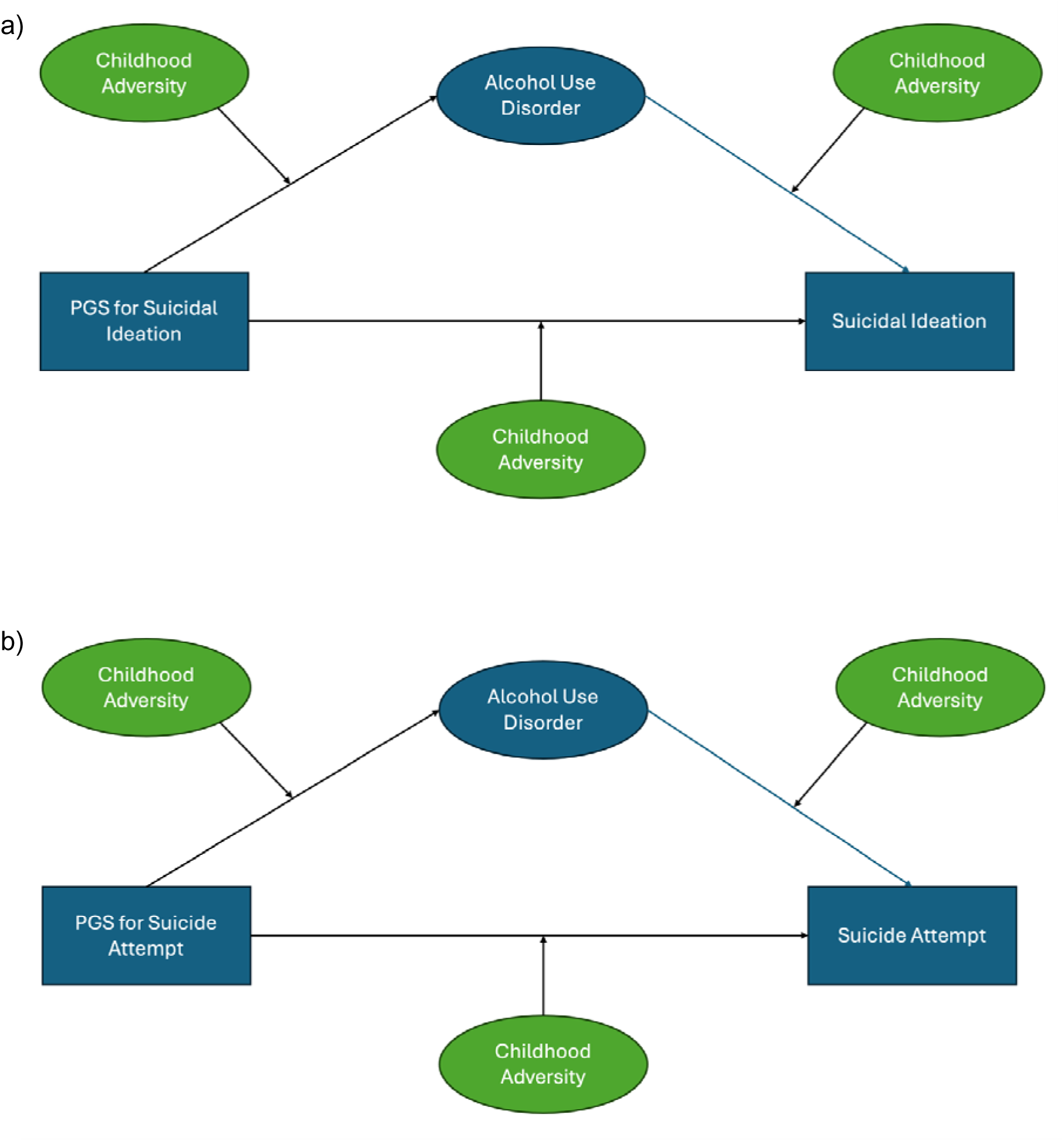
Moderated mediation model framework. *Note:* Panel A depicts the hypothesized moderated mediation model examining pathways from polygenic scores (PGS) for suicidal ideation to lifetime suicidal ideation, with alcohol use disorder (AUD) modeled as a mediator and adverse childhood experiences (ACEs) modeled as a moderator of the direct genetic pathway (PGS → suicidal ideation), the indirect pathway through AUD (PGS → AUD → suicidal ideation), and the AUD-to-outcome pathway. Panel B depicts the corresponding moderated mediation model examining pathways from PGS for suicide attempt to lifetime suicide attempt, with AUD as a mediator and ACEs as a moderator of the direct and indirect effects. In both panels, ACEs are modeled as moderators of the PGS → AUD (path a), AUD → suicidality (path b), and PGS → suicidality (path c′) associations. Models were estimated separately within African-like and European-like genetic ancestry groups and adjusted for age, sex, and ancestry principal components.

Mediation effects were estimated using quasi-Bayesian Monte Carlo simulation procedures, which repeatedly draw parameter values from the fitted models to approximate uncertainty around the estimates. We calculated the average causal mediation effect (ACME), which is the expected change in suicidality risk via AUD, and the average direct effect (ADE), which reflects the effect of PGS on suicidality after accounting for AUD. Total effects were also calculated. All effect estimates were obtained using 1,000 simulations to generate 95% confidence intervals.

To examine whether the strength of each pathway differed by ACEs exposure, we estimated moderated mediation models that included a PGS x ACEs interaction term on all three paths (a, b, and c’). This tests whether the magnitude of the direct and indirect effects varies depending on ACEs factor scores. All continuous variables were standardized within ancestry group prior to modeling to enhance interpretation, and all models included age, sex, and the first 10 ancestry PCs as covariates to reduce confounding.

To evaluate moderated mediation effects, conditional ACME values were estimated at low (−1 SD) and high (+1 SD) levels of the ACEs factor. These conditional estimates indicate whether AUD plays a stronger or weaker role in the pathway from genetic liability to suicidality depending on childhood adversity exposure. Moderation was evaluated by comparing the conditional ACME values. A false discovery rate (FDR) correction was applied across ancestry-stratified models and outcomes.

## Results

### Sample Characteristics

10,275 participants from the Yale-Penn sample were included, comprising 4,851 AFR and 5,424 EUR individuals (Table 1). Overall, 43.80% of participants were female, and the mean age was 40.59 years (standard deviation [SD] = 11.72). Lifetime SI was reported by 34.56% of participants, and 11.84% reported a lifetime SA. Participants endorsed an average of 2.44 ACEs (SD = 1.70) and 4.88 lifetime AUD criteria (SD = 4.16).

**Table 1.** Demographic and clinical characteristics of participants by genetic ancestry.

| **Phenotype** | **Full Sample**<br>**(n = 10,275)** | **AFR Individuals**<br>**(n = 4,851)** | **EUR Individuals**<br>**(n = 5,424)** |
| --- | --- | --- | --- |
|  | <u>Mean (SD) or N (%)</u> | <u>Mean (SD) or N (%)</u> | <u>Mean (SD) or N (%)</u> |
| Age | 40.59 (11.72) | 41.47 (10.16) | 39.80 (12.91) |
| Female | 4,500 (43.80) | 2,174 (44.8) | 2,326 (42.9) |
| Married | 5,846 (56.89) | 2,946 (60.73) | 2,900 (53.47) |
| Years of education | 12.78 (3.04) | 12.36 (2.74) | 13.16 (3.24) |
| Employed | 3,985 (38.78) | 1,801 (37.13) | 2,184 (40.27) |
| Suicidal ideation | 3,528 (34.56) | 1,517 (31.56) | 2,011 (37.24) |
| Suicide attempt | 1,208 (11.8) | 575 (12.0) | 633 (11.7) |
| Adverse childhood experiences | 2.44 (1.70) | 2.51 (1.70) | 2.39 (1.71) |
| Alcohol use disorder symptoms | 4.88 (4.16) | 4.74 (4.06) | 4.99 (4.24) |
| *Note: AFR = African-like genetic ancestry; EUR = European-like genetic ancestry; SD = standard deviation, N = sample size.* |  |  |  |

### Mediation Models

#### Suicidal Ideation

Direct effects of the SI PGS on SI were not significant in either ancestry group (AFR: ADE = −0.00, 95% CI [−0.02, 0.02], p = 0.99; EUR: ADE = 0.00, 95% CI [−0.01, 0.02], p = 0.55). The indirect effect of the SI PGS on SI through AUD was not significant in AFR individuals (ACME = 0.00, 95% CI [−0.00, 0.01], p = 0.32), and although statistically significant in EUR participants, the estimated magnitude was small (ACME = −0.01, 95% CI [−0.01, 0.00], p = 0.03). AUD factor scores, however, were associated with increased likelihood of SI independent of genetic risk (AFR: b = 0.54, SE = 0.04; EUR: b = 0.65, SE = 0.03; both *p*s < 2e-16).

#### Suicide Attempt

In AFR individuals, the SA PGS had a direct effect on SA after accounting for AUD (ADE = 0.36, 95% CI [0.33, 0.38], p < 2e-16). There was also an indirect effect of the PGS on SA through AUD (ACME = 0.01, 95% CI [0.01, 0.01], p < 2e-16). Similarly, in EUR individuals, the SA PGS had a direct effect on lifetime SA (ADE = 0.17, 95% CI [0.14, 0.19], p < 2e-16) and a significant indirect effect through AUD (ACME = 0.02, 95% CI [0.01, 0.02], p < 2e-16). AUD factor scores were associated with SA independent of the PGS in both ancestry groups (AFR: b = 0.53, SE = 0.07, p = 9.27e-14; EUR b = 0.71, SE = 0.06, p < 2e-16; see Figure 2).

**Figure 2.**
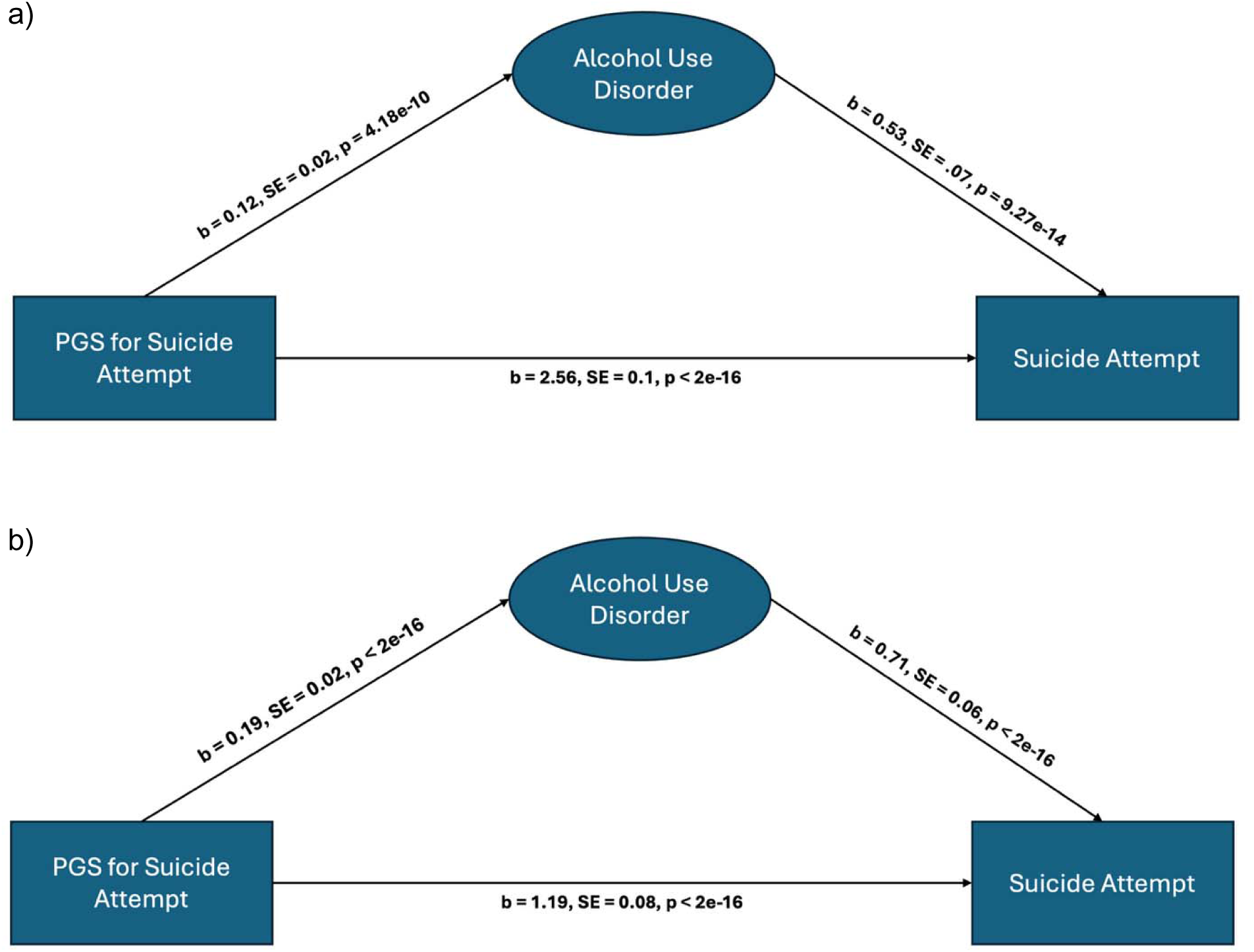
Mediation model results for suicide attempt in African-like (panel a) and European-like genetic ancestry (panel b) individuals. *Note:* Panel A depicts mediation model results for African-like genetic ancestry (AFR) individuals, showing the direct association between polygenic scores (PGS) for suicide attempt and lifetime suicide attempt, as well as the indirect pathway through alcohol use disorder (AUD). Panel B depicts corresponding mediation model results for European-like genetic ancestry (EUR) individuals. Path coefficients represent unstandardized regression estimates (b) with standard errors (SE) and two-sided P values. The direct path reflects the association between PGS and suicide attempt controlling for AUD, whereas the indirect path reflects mediation through AUD (PGS → AUD → suicide attempt). All models were adjusted for age, sex, and the first 10 ancestry principal components and were estimated separately by ancestry group.

### Moderated Mediation Models

#### Suicidal Ideation

In both ancestry groups, ACEs were positively associated with AUD (AFR: b = 0.23, SE = 0.02; EUR: b = 0.29, SE = 0.02; both *p*s < 2e-16) and SI (AFR: b = 0.66, SE = 0.05; EUR: b = 0.66, SE = 0.05; both *p*s < 2e-16). ACEs significantly moderated the AUD-SI association (path b) in AFR (interaction: b = −0.17, SE = 0.05, p = 0.001) and EUR individuals (interaction: b = −0.17, SE = 0.05, p = 0.002). In contrast, ACEs did not moderate the PGS-AUD (path a) or PGS-SI (path c′) associations (*p*s > 0.80). Consistent with this, conditional indirect effects through AUD were not significant at low or high adversity levels (AFR: ACME_low = 0.00, p = 0.66 and ACME_high = 0.00, p = 0.43; EUR: ACME_low = −0.01, p = 0.02 and ACME_high = 0.00, p = 0.98), and the difference between the ACME estimates among individuals at low and high adversity levels was not significant (AFR p = 0.86; EUR p = 0.10). Conditional direct effects (i.e., ADE) also did not differ across ACEs levels for SI (AFR: ADE_low = 0.01, 95% CI [−0.02, 0.02]; ADE_high = 0.01, 95% CI [−0.02, 0.02]; ΔADE p = 0.85; EUR: ADE_low = 0.00, 95% CI [−0.01, 0.02]; ADE_high = 0.00, 95% CI [−0.01, 0.02]; ΔADE p = 0.86).

#### Suicide Attempt

ACEs were associated with AUD in both AFR (b = 0.23, SE = 0.02, p < 2e-16) and EUR individuals (b = 0.28, SE = 0.02, p < 2e-16; see Figure 3). Additionally, ACEs were associated with SA (AFR: b = 0.61, SE = 0.09, p = 1.7e-12; EUR: b = 0.73, SE = 0.06, p < 2e-16). Moderation analyses revealed significant ACE × PGS effects on the PGS→AUD path (path a) (AFR: b = −0.05, SE = 0.02, p = 8.9e-4; EUR: b = −0.05, SE = 0.02, p = 0.006) and significant ACE × AUD effects on the AUD→SA path (path b) (AFR: b = −0.20, SE = 0.08, p = 0.01; EUR: b = −0.23, SE = 0.06, p = 0.0002). No moderation by ACEs was observed on the direct PGS→SA path (path c′) (AFR: p = 0.54; EUR: p = 0.94).

**Figure 3.**
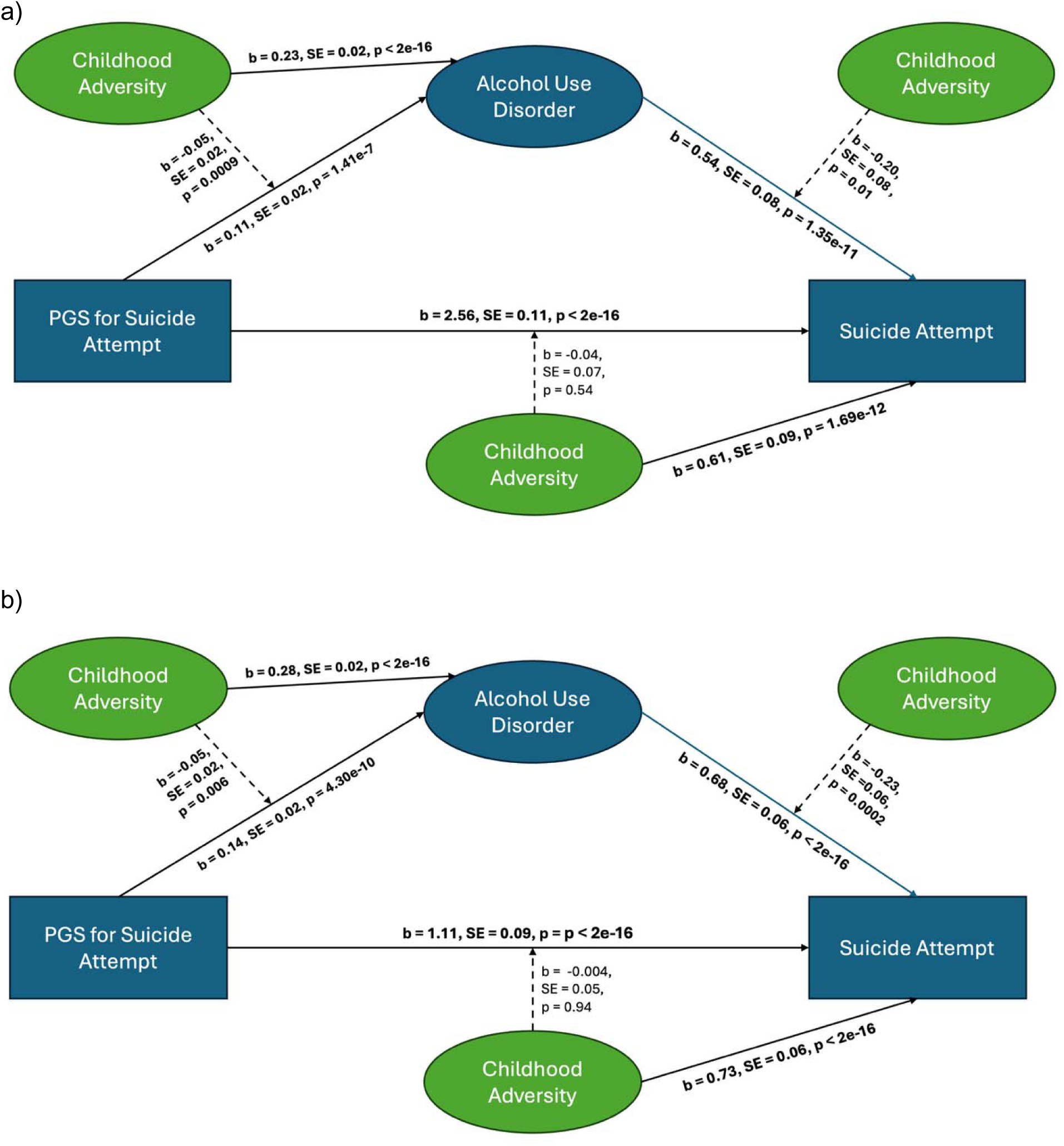
Moderated mediation results for suicide attempt in African-like (panel a) and European-like genetic ancestry (panel b) individuals. *Note:* Panel A depicts moderated mediation model results for African-like genetic ancestry (AFR) individuals, and Panel B depicts corresponding results for European-like genetic ancestry (EUR) individuals. Path coefficients shown on the arrows represent unstandardized regression estimates (b) with standard errors (SE) and two-sided P values. Solid arrows denote main effects, and dashed arrows denote interaction terms testing moderation by adverse childhood experiences (ACEs). The models include direct paths from polygenic scores (PGS) for suicide attempt to lifetime suicide attempt, indirect paths through alcohol use disorder (AUD), and moderation of the PGS → AUD (path a), AUD → suicide attempt (path b), and PGS → suicide attempt (path c′) associations by ACEs. All models were adjusted for age, sex, and the first 10 ancestry principal components and were estimated separately by ancestry group.

Conditional indirect effects differed across levels of ACEs. In AFR individuals, the ACME estimate was larger among individuals with low adversity (ACME_low = 0.01, p < 2e-16) than high adversity (ACME_high = 0.00, p = 0.04; ΔACME = 0.02, p < 2e-16). A similar pattern was observed in EUR individuals, with significant ACME estimates at both levels of ACEs (ACME_low = 0.01, p < 2 × 10_−1_□; ACME_high = 0.01, p = 0.002) and a larger indirect effect among individuals with low adversity (ΔACME = 0.01, p = 0.03). Conditional direct effects also varied by ACEs level. In AFR individuals, ADE estimates increased from low to high adversity (ADE_low = 0.26, 95% CI [0.22, 0.30]; ADE_high = 0.40, 95% CI [0.36, 0.43]; ΔADE p < 2e-16). A similar pattern was observed in EUR individuals (ADE_low = 0.08, 95% CI [0.06, 0.11]; ADE_high = 0.20, 95% CI [0.17, 0.24]; ΔADE p < 2e-16).

## Discussion

This study examined how genetic liability, AUD, and childhood adversity jointly contribute to SI and SA in AFR and EUR individuals. Three key findings emerged. First, polygenic liability for SA had direct and indirect effects on SA, whereas SI PGS had no such effects on SI. Second, AUD partially mediated the association between genetic liability and SA in both AFR and EUR individuals, indicating that alcohol misuse may be one behavioral mechanism through which inherited vulnerability to suicidality manifests. However, the magnitude of these indirect effects was modest, accounting for ∼2% of the total effect in AFR individuals and ∼10% in EUR individuals. Third, ACEs moderated these pathways for SA, such that indirect effects of PGS through AUD were reduced at higher adversity levels. In contrast, no moderated mediation was observed for SI. Together, these findings highlight heterogeneity in the etiological pathways that underlie suicidality and point to an alcohol-related pathway to SA that is conditional on early environmental exposures.

Consistent with our hypotheses, AUD emerged as a significant mediator of genetic risk for SA. These findings align with a large body of research indicating that alcohol use is one of the most potent proximal risk factors for suicidal behavior, reflecting its effects on impulsivity, judgment, and exposure to lethal means.^11–15^ Although the direct effects of PGS on suicidality were small, the presence of significant mediation suggests that genetic liability to suicidality may be expressed partly through alcohol-related mechanisms.^13,30,41^ Notably, mediation was stronger for SA than SI, supporting theoretical models in which alcohol’s effects on impulsivity and disinhibition may be especially critical for facilitating the transition from suicidal thoughts to attempts. These models emphasize the impaired self-control characteristic of alcohol use as a driver of suicidal behavior.^5,13^ Clinically, these findings suggest that targeting alcohol misuse may reduce risk for SA among individuals at high genetic liability and highlight the importance of incorporating alcohol use screening and interventions into suicide prevention frameworks.

Under a diathesis-stress model, childhood adversity should amplify the translation of genetic risk into behavioral outcomes.^28,29^ In contrast, we found that the indirect effects of polygenic liability on suicidality through AUD were attenuated at higher adversity levels in both ancestry groups. Thus, among individuals with greater ACEs exposure, suicidality may emerge primarily through pathways less dependent on alcohol, including trauma-related psychopathology, emotion dysregulation, or stress reactivity.^23,26,30,35,42^ Under conditions of lower childhood adversity, however, alcohol involvement appeared to play a more prominent role in translating genetic vulnerability into suicidal behaviors. This pattern is consistent with multifinality models in which suicidality can arise through multiple etiological pathways (e.g., an alcohol-related disinhibitory pathway and a trauma-related affective pathway that diminishes the explanatory contribution of alcohol). These findings may also have relevance for personalized intervention approaches. Individuals with substantial childhood adversity may benefit most from trauma-focused approaches targeting dysregulated affect and interpersonal stress, whereas individuals with lower childhood adversity may derive greater benefit from interventions directly targeting alcohol misuse.

## Limitations

Several limitations warrant consideration. First, PGS were derived from GWAS of SI and SA that had substantially larger EUR than AFR discovery samples, reflecting broader inequities in genetic research. Despite this difference in statistical power, we observed consistent effects in the two ancestry groups: the SA PGS significantly predicted SA in both AFR and EUR individuals, while the SI PGS failed to predict SI in either group. This pattern suggests that the lack of association for SI is less likely to be attributable to ancestry-related differences in statistical power. Future work leveraging larger and more ancestrally diverse GWAS samples may nonetheless improve the precision and generalizability of PGS-based analyses. Second, although we modeled ACEs as a latent factor, specific types of ACEs (e.g., physical abuse vs. neglect) may differentially influence suicidality, and this possibility warrants examination in future research.^35^ Third, the Yale-Penn sample includes only cross-sectional data, precluding causal inference from the mediation models. Although this is partially alleviated by PGS, which are determined prior to alcohol use and suicidality, longitudinal designs are needed to validate findings.

## Conclusions

By integrating genetic, behavioral, and environmental risk factors in moderated mediation models within EUR and AFR individuals, this study advances our understanding of the pathways to the development of suicidality. The findings identify alcohol involvement as a partial behavioral mediator of genetic liability to SA and show that this mechanism is more relevant in the context of lower childhood adversity. These results underscore the importance of considering multiple etiological pathways to suicidality and the need for precision medicine approaches tailored to an individual’s genetic, behavioral, and environmental risk profiles to reduce the global burden of suicide.

## Data Availability

Analysis code is available upon reasonable request to the authors. The individual level Yale-Penn data are not available due to the confidentiality procedures of the study.

## Notes

**Funding:** This research was supported by the Department of Veterans Affairs Office of Academic Affiliations Advanced Fellowship Program in Mental Illness Research and Treatment, the Crescenz VA Mental Illness Research, Education, and Clinical Center, and the National Institute of Mental Health (R01MH133728). Dr. Kimbrel was supported by a VA Research Career Scientist (RCS) Award (I01BX005881) from the Biomedical and Laboratory Research Service of the U.S. Department of Veterans Affairs Office of Research and Development. The views expressed in this article are those of the authors and do not necessarily reflect the position or policy of the VA, the U.S. government, Duke University, or any other affiliated institution.

### Competing Interest Statement

Dr. Kranzler is a member of advisory boards for Altimmune and Clearmind Medicine; a consultant to Sobrera Pharmaceuticals, Altimmune, Lilly, and Ribocure; and the recipient of research funding and medication supplies for an investigator-initiated study from Alkermes and company-initiated studies by Altimmune and Lilly. Dr. Gelernter is paid for editorial work for the journal Complex Psychiatry. All other authors declare no conflicts of interest.

### Funding Statement

This research was supported by the Department of Veterans Affairs Office of Academic Affiliations Advanced Fellowship Program in Mental Illness Research and Treatment, the Crescenz VA Mental Illness Research, Education, and Clinical Center, and the National Institute of Mental Health (R01MH133728). Dr. Kimbrel was supported by a VA Research Career Scientist (RCS) Award (I01BX005881) from the Biomedical and Laboratory Research Service of the U.S. Department of Veterans Affairs Office of Research and Development. The views expressed in this article are those of the authors and do not necessarily reflect the position or policy of the VA, the U.S. government, Duke University, or any other affiliated institution.

### Author Declarations

This study was approved by the Institutional Review Board of the University of Pennsylvania (protocol #804787 and #812856).

