## Supplementary Tables for "Pathways from Polygenic Risk to Suicidality: Effects of Alcohol Use Disorder and Childhood Adversity"

| Fit Index | Value |
| --- | --- |
| χ² (df) | 1627.57 (44) |
| P value (χ²) | < 0.001 |
| Comparative Fit Index (CFI) | 0.950 |
| Tucker–Lewis Index (TLI) | 0.938 |
| Root Mean Square Error of Approximation (RMSEA) | 0.068 |
| 90% CI for RMSEA | 0.065–0.071 |
| P value (RMSEA ≤ 0.05) | < 0.001 |
| P value (RMSEA ≥ 0.08) | < 0.001 |
| Standardized Root Mean Square Residual (SRMR) | 0.034 |
| Akaike Information Criterion (AIC) | 82,820.05 |
| Bayesian Information Criterion (BIC) | 82,972.97 |
| Sample-size adjusted BIC (SABIC) | 82,903.06 |

Supplementary Table S1. Model fit indices for the alcohol use disorder (AUD) latent factor.

*Note:* Alcohol use disorder (AUD) was modeled as a single latent factor based on 11 DSM-5 AUD criteria. Model fit indices indicated good to acceptable fit (CFI = 0.95; TLI = 0.94; RMSEA = 0.07; SRMR = 0.03). Factor scores derived from this model were standardized within genetic ancestry group and used as the mediator in all analyses.

Supplementary Table S2. Model fit indices for the adverse childhood experiences (ACEs) latent factor.

| Fit Index | Value |
| --- | --- |
| χ² (df) | 473.96 (34) |
| P value (χ²) | < 0.001 |
| Comparative Fit Index (CFI) | 0.915 |
| Tucker–Lewis Index (TLI) | 0.887 |
| Root Mean Square Error of Approximation (RMSEA) | 0.036 |
| 90% CI for RMSEA | 0.033–0.039 |
| P value (RMSEA ≤ 0.05) | 1.000 |
| P value (RMSEA ≥ 0.08) | < 0.001 |
| Standardized Root Mean Square Residual (SRMR) | 0.030 |
| Akaike Information Criterion (AIC) | 315,121.09 |
| Bayesian Information Criterion (BIC) | 315,344.21 |
| Sample-size adjusted BIC (SABIC) | 315,245.70 |

*Note:* Adverse childhood experiences (ACEs) were modeled as a single latent factor using ten dichotomized indicators reflecting childhood adversity or absence of protective factors before age 13. Model fit indices indicated good fit (CFI = 0.92; RMSEA = 0.04; SRMR = 0.03). Residuals for household substance use and household smoking were allowed to covary to improve model fit. Factor scores derived from this model were standardized within genetic ancestry group and used as moderators in subsequent analyses.
